# What is the relationship between viral prospecting in animals and medical countermeasure development?

**DOI:** 10.1101/2024.08.09.24311747

**Authors:** Aishani V. Aatresh, Marc Lipsitch

## Abstract

In recent decades, surveillance in nonhuman animals has aimed to detect novel viruses before they “spill over” to humans. However, the extent to which these viral prospecting efforts have enhanced preparedness for disease outbreaks remains poorly characterized, especially in terms of whether they are necessary, sufficient, or feasible ways to spur medical countermeasure development. We find that several viruses which pose known threats to human health lack approved vaccines and that known viruses discovered in human patients prior to 2000 have caused most major 21^st^-century outbreaks. With *Filoviridae* as a case study, we show there is little evidence to suggest that viral prospecting has accelerated countermeasure development or that systematically discovering novel zoonotic viruses in animal hosts before they cause human outbreaks has been feasible. These results suggest that prospecting for novel viral targets does not accelerate a rate-limiting step in countermeasure development and underscore questions about the importance of zoonotic viral discovery for outbreak preparedness. We consider limitations to these conclusions and alternative but related approaches to preparedness and response.

## Introduction

In recent decades, concerns about viruses that have the potential to “spill over” from non-human animals to humans and cause the next infectious disease outbreak or widespread epidemic have driven systematic efforts to characterize viral diversity and discover novel viral species in animal hosts, including those that may be or become reservoirs for spillover into humans.^1, 2, 3, 4^ The COVID-19 pandemic has intensified a focus on and controversies around attempts at viral discovery as a means to isolate potentially zoonotic or currently unknown “Disease X” viruses from animals before they spill over to humans.^5^ Surveillance in animals, especially wildlife, to this end has been a central element of One Health initiatives in renewed discussions about and global efforts toward pandemic prevention, preparedness, response and resilience.^6, 7^ However, some elements of animal virus surveillance projects remain poorly defined, namely: how knowledge of viral zoonoses in animals has contributed to outbreak preparedness and response, what assumptions motivate large-scale viral surveillance in animals and whether they hold, and whether the benefits claimed to be attainable through such viral discovery have proven realistic.

Current estimates suggest that approximately 250 viruses are known to infect humans^8^ and implicate zoonotic spillovers as the proximate causes for approximately 60% of emerging infectious disease events, with wildlife origins for about 70% of these spillovers.^9, 10^ Examples of such emerging infectious disease events — defined as the first occurrence of a pathogen in a human population, an increase in the geographic range or incidence of a previously known pathogen, or a major change in pathological or clinical presentation — range from Crimean-Congo hemorrhagic fever (CCHF) and Nipah virus infection to yellow fever and Ebola virus disease. Remaining diseases have emerged or were caused by other types of pathogenic agents (e.g., bacteria or fungi), human-to-human transmission of a virus, or a pathogen of unknown origin. One study posits that over 600,000 unknown viruses with the potential to infect humans lurk in birds and non-human mammals,^11^ while another more recent estimate suggests that 10,000 viruses with zoonotic potential circulate in non-human mammals.^12^

The extent of unknown viral diversity and possible opportunities to better understand viral ecology and disease emergence have been invoked to champion discovering viruses in animal hosts before human outbreaks as a means to improve global preparedness for such outbreaks. PREDICT is one notable example of a viral discovery program with a focus on systematically sampling animals for viruses of future relevance. It was led by the United States Agency for International Development (USAID) and University of California at Davis from 2009 through 2020 with partnerships in over 30 countries. This project focused on “[p]redicting where new diseases may emerge from wild animals, and detecting other pathogens before they spread among people [to] give us the best chance to prevent new pandemics”^13^ by advancing an “approach in which pathogens of pandemic potential are discovered at their source before large-scale epidemics occur in people.”^1^ We term these systematic predictive efforts for “finding viruses in wildlife before they emerge in humans”^14^ *viral prospecting* to distinguish discovering and forecasting future disease threats from capacity-building for surveillance of known viruses, another major aim of PREDICT-style programs.

To date, assessments of animal viral discovery efforts primarily have focused on the distribution of and disease burden in host and reservoir species,^4, 15^ sampling and viral discovery curves, ^16, 17, 18^ cost-benefit analyses,^19^ and predictive methods to compute the likelihood of spillover across viral species given various variables of interest.^20, 21^ Efforts to rank which known viruses are most likely to spill over from animals to humans have incorporated novel sequences from such efforts with expert predictions and ecological data. One such study suggests that the top 12 are zoonotic viruses that have already been well-characterized, while half of the remaining top 50 were various coronaviruses sampled primarily from bats. Notably, this study used observed spillover as one of its predictors, risking circularity.^22^

Critiques of virus prospecting have revolved around safety risks and, to a lesser extent, whether the viruses discovered are of significant value for human health. Some reports in the literature document wildlife biologists bitten by host species known to harbor viral zoonoses during field sampling efforts, with infection in some cases.^23, 24^ With more recent controversies around the origins of the COVID-19 pandemic, biosafety has also been raised as reason to terminate large-scale prospecting projects.^25, 26, 27^ Other commentaries focus on mechanisms to mitigate zoonotic spillover^7, 28^ or concerns about the feasibility of prospecting efforts and its consequences of unmet expectations for trust in science.^29^ For instance, PREDICT detected over 800 novel viruses, some of which were novel strains of known viruses based on a pairwise sequence identity cut-off, in samples from approximately 7,000 animals tested across 15 taxonomic families with known viruses that infect humans.^22^ However, project meta-analyses note that out of the total effort — more than 500,000 samples primarily from over 70,000 animals — just one of these novel viral species is known to be capable of causing disease in humans, and it was only discovered while research teams were supporting the investigation of an outbreak in humans.^30, 31^

The promises of viral prospecting efforts like PREDICT extend beyond advances in basic science and knowledge of ecological diversity or international development. Some researchers have argued that viral prospecting holds direct benefits for predicting disease emergence and accelerating medical countermeasure (MCM) development for those future diseases.^32, 33, 34, 35, 36, 37, 38^ Having vaccines at-the-ready during an emerging outbreak has become an increasingly central focus in disease preparedness discussions and predictive efforts over the past two decades, with the goal that such development of medical countermeasures prior to possible large-scale human outbreaks will reduce their morbidity, mortality, and economic consequences.^39, 40, 41^ The literature to date pays comparatively less attention to evidence for these potential translational impacts of viral discovery. Nonetheless, next-generation viral prospecting projects call for over one billion dollars in funding and “envisage countries working together to fund viral discovery programs that upload sequence data in almost real-time, so that it can be used to identify those microbes most likely to be able to cause zoonoses, and the data then can be used to block spillover and create vaccines.”^42^

Put differently, rationales for surveillance projects to predict future outbreaks advance and are supported by the potential for specific follow-on interventions and benefits from them. These include building technical capacity in areas of high spillover potential or reducing contact at human-wildlife interfaces to contain viruses — for example, by measures that restrict live animal sales or hunting reservoir species. However, this Analysis approaches viral prospecting in terms of aforementioned MCM development-related rationales, which is one of the most tangible and widely discussed objectives for 21^st^-century preparedness efforts. We assess to what extent viral prospecting especially in wildlife has enabled accelerated medical countermeasure development to strengthen preparedness and response for past outbreaks in humans. Where other studies have aimed to weigh tradeoffs and risks or costs and benefits of animal surveillance, we focus on first understanding these possible benefits: what would need to be true for them to be realized as suggested, and to what extent have foundational assumptions and projections about them materialized over time?

In light of calls for intensified animal surveillance to predict and prepare for future epidemics, we attempt to address these questions by testing two primary hypotheses: that viral surveillance in animals identifies potential (pre-emptive) and actual (post-hoc) causes of outbreaks in humans, and that it translates to accelerated MCM development. We identify and assess a series of related conditions that would need to hold true to justify arguments for the necessity, sufficiency, and feasibility of animal viral prospecting: (i) past efforts at viral prospecting have paid off in that scientists have first discovered in zoonotic hosts viruses which do later cause disease in humans; (ii) the discovery of a virus in an animal host prior to the first outbreak in humans has translated to enhanced preparedness and response capabilities, defined primarily through available MCMs; (iii) novel viruses are causing outbreaks and plausibly could have been discovered in an animal host prior to these outbreaks; (iv) viruses prioritized for MCM development were discovered in animal hosts; and (v) viral prospecting supplies needed candidates for MCM development because efforts to deal with all or most viral threats are underway and succeeding. We evaluate these hypotheses and assumptions through a series of case studies across virus families, and we consider limitations of these analyses alongside possible alternative approaches.

## Results

### Discovery of a virus prior to its first documented outbreaks in humans has limited bearing on preparedness and response

We first evaluated which viruses known to cause disease in humans were first discovered in animals and whether their discovery in an animal host or vector has had a substantial bearing on preparedness and response. In assessing how each of the approximately 250 viruses known to infect humans was first isolated, we identified eleven viruses that were isolated in animals prior to causing eventual clusters of cases in humans (Table 1). With the exception of monkeypox and Puumala viruses, all of these viruses are transmitted by mosquitoes and cause vector-borne diseases. Three — monkeypox, Rift Valley Fever, and Zika — have caused several notable outbreaks of disease in humans in the decades since their discovery. Knowledge of these viruses from an animal source prior to their first outbreaks in humans has not translated to a distinctively robust capacity to prevent or respond to future outbreaks; each has spread beyond previously contained regions.^43, 44, 45^ Furthermore, there is no licensed vaccine against Zika or Rift Valley Fever for use in humans, while those approved for monkeypox virus are at the time of writing all repurposed or expanded-use smallpox vaccines.^46^ The other viruses on this list cause seemingly limited disease in humans as detected through often-scant diagnostic and surveillance capacity.

**Table 1:** Viruses first discovered in animals before causing an outbreak in humans.

| Virus | Discovered | 1<sup>st</sup> documented human outbreak | Conditions of isolation and description |
| --- | --- | --- | --- |
| Barmah Forest | 1989 | 1992 | *Culex* mosquitoes in Australia |
| Bunyamwera | 1943 | 1955 | Through yellow fever surveillance in Uganda |
| Eastern equine encephalitis | 1831 | 1938 | North American horses |
| Monkeypox | 1958 | 1970 | Research monkeys; first observed human infection in child in DRC. Zoonotic reservoir unknown. |
| Ngari | 1979 | 1993 | *Aedes simpsoni* mosquitoes in Senegal |
| Puumala | 1979 | 2000 | Bank voles in Finland |
| Rift Valley Fever | 1931 | 1975 | Outbreak on a sheep farm in Kenya |
| Semliki Forest | 1942 | 1987 | *Aedes abnormalis* mosquitoes in western Uganda |
| Sindbis | 1952 | 1961 | *Culex* mosquitoes in the Nile River Delta, Egypt; used as a model alphavirus in USA |
| Venezuelan equine encephalitis | 1938 | 1950 | From the brain of a deceased horse |
| Zika | 1947 | 1952 | Through yellow fever surveillance in infected rhesus macaques for research use |

### 21st century outbreaks of international concern largely have been caused by known viruses first discovered in humans

We next assessed whether other previously novel viruses, which might have plausibly been discovered in animals, are causing high-consequence outbreaks in humans. Data from the World Health Organization’s (WHO) Disease Outbreak News system (DON) and Public Health Emergency of International Concern (PHEIC) declarations both suggest otherwise. Since 1996, DON has function as the WHO’s public online system to compile and disseminate disease event reports from countries and partner organizations. We subset Carlson et al.’s 1996-2019 DON database ^47^ for viral disease events to analyze where and which viruses are causing outbreaks. We find that while certain regions dominate DON reports, a limited number of known viruses are causing the majority of internationally reported disease events worldwide (Figure 1). Across regions, the plurality of DONs are concentrated around Ebola virus, Influenza A, MERS-CoV, and Yellow fever. This distribution largely reflects extended and widespread outbreaks of known viruses (e.g., Ebola) or spikes in endemic disease (e.g., Yellow fever), while fewer DONs report instances in which spillover of a novel virus (e.g., initial emergence of MERS-CoV, Nipah virus) drives disease.

**Figure 1:**
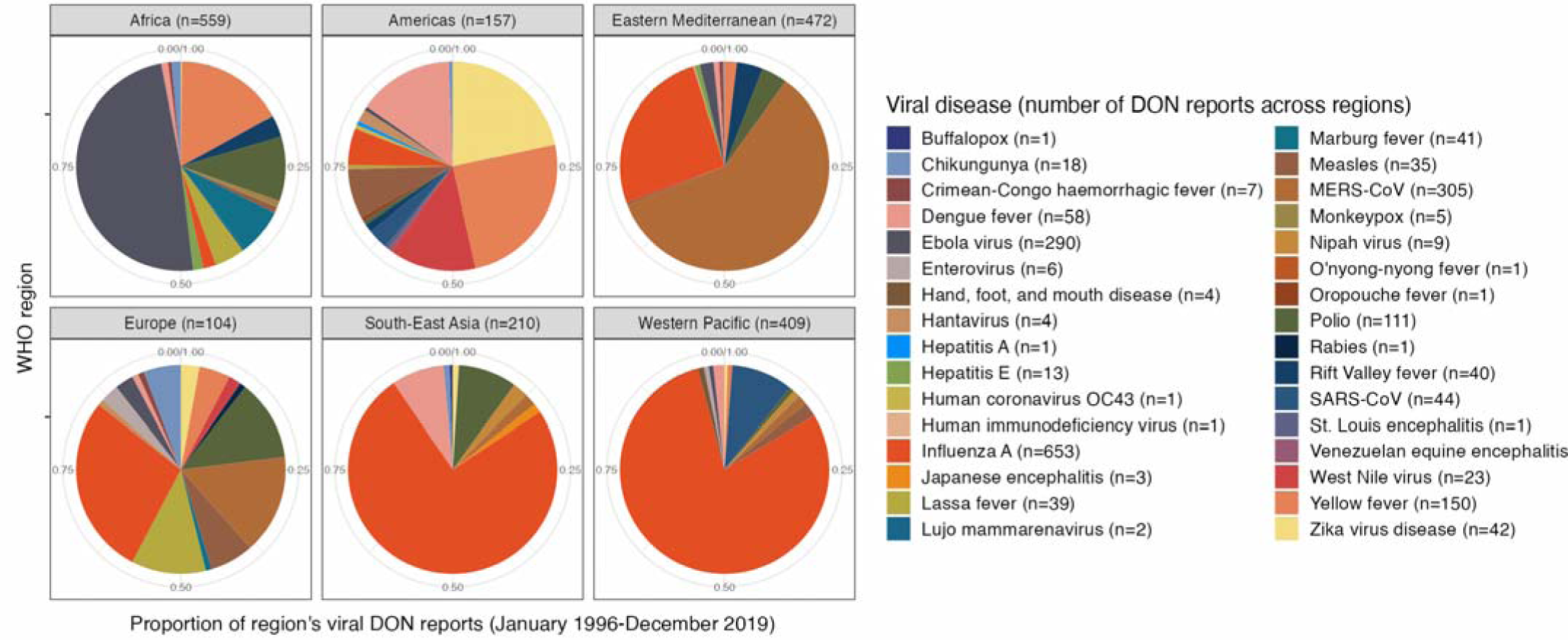
Geographic and viral distribution of WHO DON reports (1996-2019) Regional distribution for virus-only subset of WHO Disease Outbreak News reports from Carlson et al. (2023) database.

DON mirrors countries’ infectious disease concerns, but reports primarily offer weekly updates and early warning signals during an emerging outbreak rather than definitive markers of escalating disease emergencies. PHEIC declarations, on the other hand, reflect assessments that a disease event poses a severe global threat.^48^ The WHO has declared 7 PHEICs since the instrument was established under the 2005 revision to the International Health Regulations, following the 2002-2004 SARS epidemic (Table 2). PHEICs primarily have been associated with viruses that were well-characterized or that had caused outbreaks in humans prior to the disease event in question. In the two events (H1N1 and COVID-19) of a novel viral strain or species leading to a PHEIC, the pathogen nonetheless belonged to an extensively studied virus family. These data suggest that spillovers of novel zoonotic pathogens are not driving outbreaks that countries report to the WHO or that international public health experts deem most emergent.

**Table 2:** WHO PHEIC declarations by viral disease and discovery status.

| Year | Viral Disease | Novel Pathogen? |
| --- | --- | --- |
| 2009 | H1N1 | TRUE |
| 2014 | Ebola |  |
| 2014 | Polio (*ongoing PHEIC*) |  |
| 2016 | Zika |  |
| 2018 | Ebola |  |
| 2020 | COVID-19 | TRUE |
| 2022 | Mpox |  |

### Viral prospecting makes limited contributions to MCM development pipelines

The analyses described in the preceding sections characterized what viruses are causing disease in humans and the relationship they have to viruses discovered in animal hosts over time. We next sought to address whether viral discovery in animal hosts addresses a rate-limiting step in overall MCM development efforts, with a specific focus on vaccines as primary instruments for preventing infection and the severity or spread of disease. Specifically, we aimed to determine if viral prospecting has discovered pathogens of interest for global efforts to develop MCMs against epidemic threats and whether scientists lack pathogens worth targeting to these ends, such that viral prospecting might be necessary to expand vaccine development horizons.

Since 2018, comprehensive priority lists for MCM development have been released by four institutions: the Coalition for Epidemic Preparedness Innovations (CEPI),^49^ U.S. National Institute of Allergy and Infectious Diseases (NIAID),^50^ U.K. Vaccine Network (UKVN),^51^ and WHO.^52^ We find that viruses prioritized for MCM development by at least one of these agencies were predominantly first isolated well before 2000 and during the first documented outbreak of each virus in humans (Figure 2). Systematic viral prospecting efforts from the 21^st^ century have not contributed any novel zoonotic viruses to these lists, and surveillance in animals more broadly has played a limited role in isolating priority viruses.

**Figure 2:**
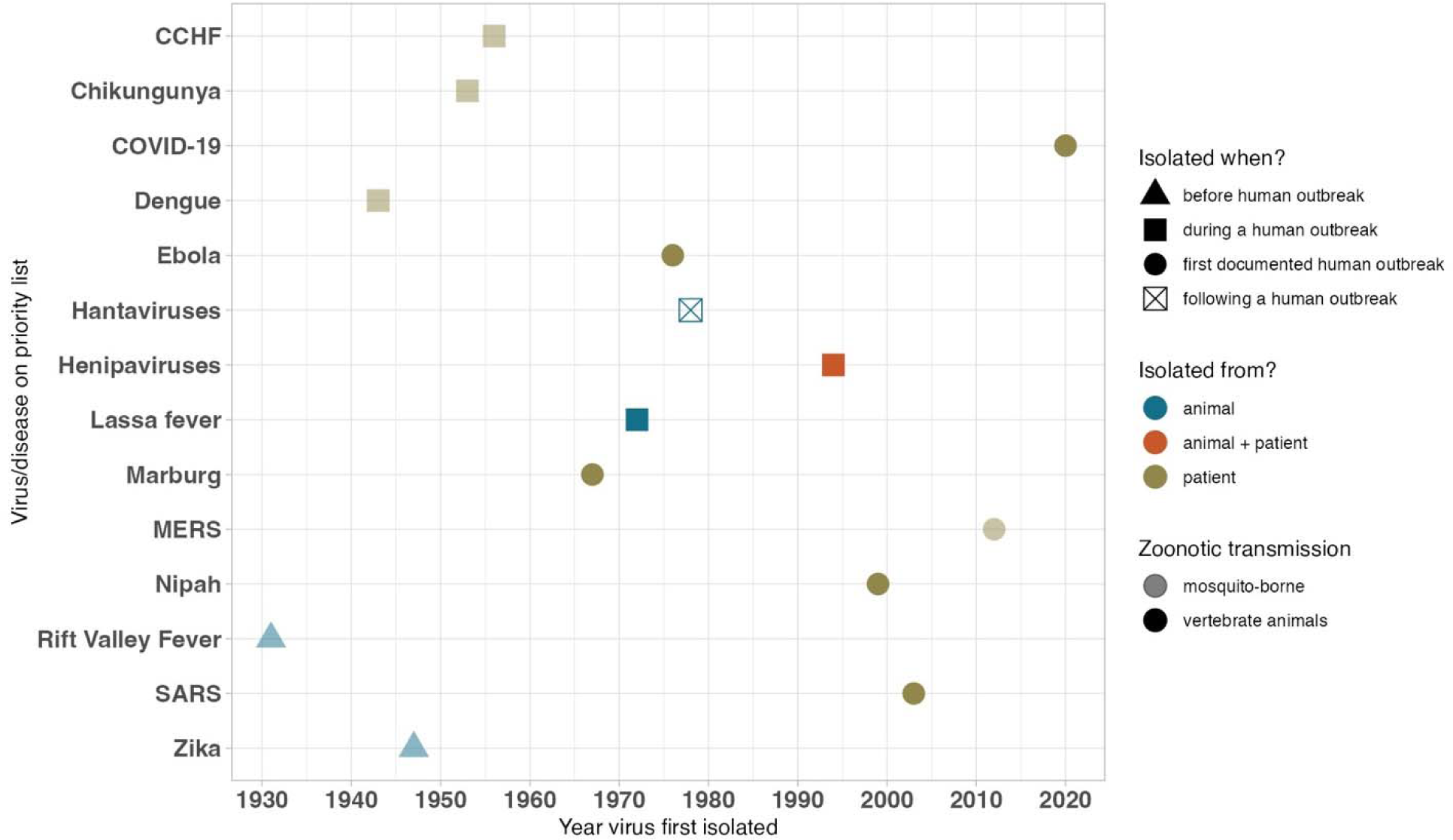
Characteristics of viral pathogens on priority lists for MCM research and development. Priority pathogens for research under WHO, NIAID (Category A), UKVN, and/or CEPI were assessed for mode of transmission, first recorded outbreak, and circumstances under which they were first isolated.

Next, we characterized the state of vaccine development against known viral threats. Many vaccine development efforts in the wake of COVID-19 and other recent epidemics have been schematized taxonomically by virus family, of which by some classifications there are 26 with at least one virus known to infect humans.^53^ We find that the first virus isolated in the majority of these families was discovered in humans between 1940 and 1980 (Table 3). At present, a regulatory body has approved at least one vaccine for use in humans against at least one virus in 16 of these families. Prior to 2020, this number was 14. The first vaccine approved for a coronavirus was against SARS-CoV-2 during the COVID-19 pandemic, even though this vaccine relied on years of research and development efforts against SARS-CoV and MERS-CoV following human outbreaks of these two coronaviruses. The first vaccine for a pneumovirus was approved in 2023 and targets respiratory syncytial virus (RSV). Of the 10 known families for which no vaccine is currently approved for any virus in each family, there are some striking absences. The lack of a human vaccine for any retrovirus or bunyavirus is notable, for example; HIV is a prominent retrovirus that has afflicted humans for decades, and Rift Valley Fever, Crimean-Congo hemorrhagic fever, and hantaviruses are long-standing disease threats from the bunyavirus family. In summary, significant gaps remain in vaccine development against known targets, many of which like HIV have been the focus of widespread efforts and posed significant technical challenges. Zoonotic viruses discovered in animal hosts have contributed few previously unknown targets of interest to further direct these efforts.

**Table 3:** Vaccine development and viral discovery across virus families.

| *Family* | *Year 1<sup>st</sup> vaccine approved for human use* | *Year 1<sup>st</sup> virus isolated, source* | *Notes* | *Examples of notable viruses* |
| --- | --- | --- | --- | --- |
| Adenoviridae | 1971 | 1953, human | Only vaccine against an adenovirus approved for use in US military; separate from adenoviruses used as vectors for vaccines against other pathogens |  |
| Anelloviridae | -- | 1997, human | Estimated >90% prevalence in population, primarily nonpathogenic |  |
| Arenaviridae | 2006 | 1933, human | 2006 Junin virus vaccine, only licensed for use in Argentina | Lassa fever |
| Astroviridae | -- | 1975, human |  |  |
| Bornaviridae | -- | 1975, animal | Horses and sheep extensively vaccinated |  |
| Bunyavirales | -- | 1931, animal | Veterinary vaccine used extensively in Africa for Rift Valley Fever (RVF) | RVF, CCHF, Hantavirus |
| Caliciviridae | -- | 1972, human | Cats vaccinated; no human concerns beyond norovirus |  |
| Coronaviridae | 2020 | 1965, human |  | SARS-CoV-2, SARS-CoV-, MERS-CoV |
| Filoviridae | 2019 | 1967, human |  | Ebola, Marburg |
| Flaviviridae | 1937 | 1927, human |  | Dengue, Zika, Japanese encephalitis, Yellow fever |
| Hepadnaviridae | 1981 | 1965, human |  | Hep B |
| Hepeviridae | 2011 | 1990, human | Vaccine licensed only in China | Hep E |
| Herpesviridae | -- | 1919, human |  |  |
| Orthomyxoviridae | 1945 | 1933, human |  | Influenza |
| Papillomaviridae | 2006 | 1979, human |  | HPV |
| Paramyxoviridae | 1963 | 1934, human |  | Measles, mumps, Nipah |
| Parvoviridae | -- | 1960, animal | Some disease in humans but usually asymptomatic; human trials stopped for severe adverse events. Family established 1975. Human disease first observed 1974. |  |
| Picobirnaviridae | -- | 1988, human | Opportunistic enteric pathogens |  |
| Picornaviridae | 1955 | 1898, animal |  | Polio |
| Pneumoviridae | 2023 | 1956, animal | Separated from<br>*Paramyxoviridae*, 2016 | RSV |
| Polyomaviridae | -- | 1953, animal |  |  |
| Poxviridae | 1796 | -- | Edward Jenner and cowpox-<br>based vaccination for<br>smallpox | Smallpox,<br>monkeypox |
| Reoviridae | 1998 | 1953, human |  | Rotavirus |
| Retroviridae | -- | 1908, animal |  | HIV |
| Rhabdoviridae | 1885 | -- | Louis Pasteur and inactivated<br>rabies virus vaccine | Rabies |
| Togaviridae | 1969 | 1930, animal | Separated from *Flaviviridae*,<br>1984 | Rubella |

### Viral discovery in animals plays a limited role in MCM development and outbreak response for filoviruses that infect humans

Finally, we aimed to further specify relationships between animal viral discovery and MCM development through a case study of the *Filoviridae* family. Two of its genera, *Ebolavirus* and *Marburgvirus*, include zoonotic viral species that have caused several outbreaks in humans over the past 50 years. The 2013-2016 Ebola epidemic in West Africa is the largest filovirus outbreak to date, with over 28,000 cases and 11,000 deaths.^54^ Since the first international reports of disease several decades ago, extensive resources have been devoted to detecting filoviruses in animals. Outbreaks of both Ebola virus disease (EVD) and Marburg virus disease (MVD) in 2022 and 2023 notably occurred in countries across sub-Saharan Africa where cases had not been previously documented, underscoring increasing international concern about filoviruses.

The first filovirus cases in humans were documented in 1967 during two simultaneous Marburg virus outbreaks in Germany and then-Yugoslavia, both of which were traced back to infected laboratory African green monkeys (Figure 3A). The next several Marburg cases were predominantly reported in tourists who had visited caves that were known bat habitats in various African countries’ national parks, with occasional reports of disease in miners. Other outbreak investigations have found either tenuous or no epidemiological or genomic links to bats, in caves or elsewhere, as a putative source of infection (Supplementary Information). The largest MVD outbreak to date began in Angola in 2004, infecting over 250 people and causing more than 225 deaths. It was preceded by an outbreak in 1998 in the Democratic Republic of the Congo (DRC), now the second-largest Marburg outbreak, during which epidemiological investigations revealed several possible years of internationally unreported MVD.^55^ As early as 1987, local doctors and two hospitals had been treating patients with “a disease called ‘syndrome hémorragique de Durba,’ which was always associated with mining, was common knowledge among villagers and health care workers in the area” and caused several outbreaks of more than 50 individuals documented only in local records.

**Figure 3:**
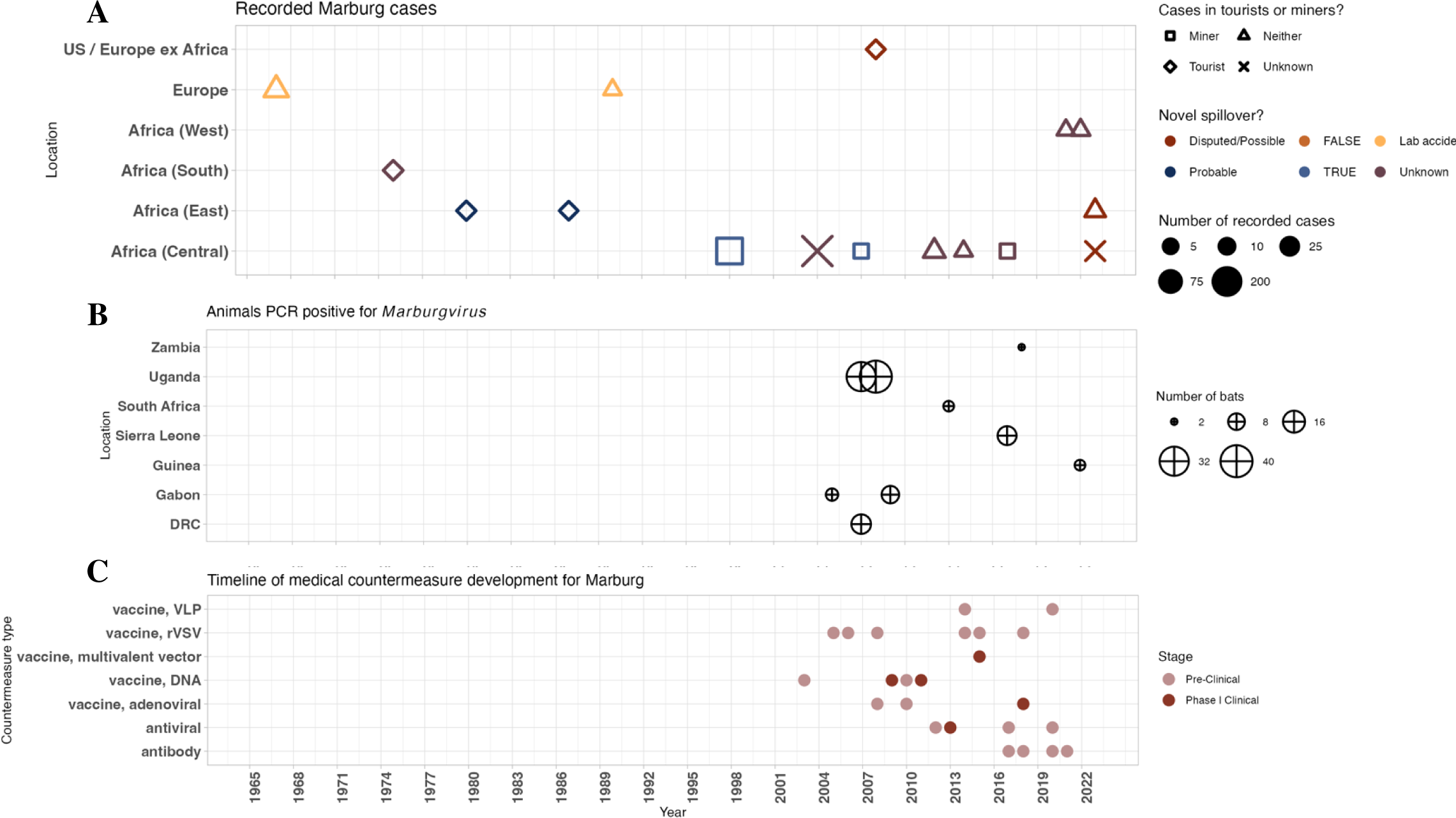
Marburg outbreaks, animal surveillance, and MCM development. A) Timeline of MVD outbreaks by country, assessment of novel spillover event as source of outbreak, and primary outbreak population. The Angola outbreak, the largest recorded, is the large “X” at 2004 and Africa (Central). B) Timeline of when and where samples from animals were positive for Marburg virus by polymerase chain reaction. C) Timeline of published pre-clinical (and non-murine) work and clinical trials by type of medical countermeasure specifically targeting Marburg virus.

In 2007, animal surveillance definitively established Egyptian fruit bats as a reservoir host for Marburg by isolating live virus from several animals (Figure 3B). This discovery was preceded by 16 samples from bats in caves from the DRC and Gabon that were positive for Marburg by polymerase chain reaction (PCR) and contemporaneous with 40 samples PCR positive for a *Marburgvirus* obtained from August 2008 through December 2009. These discoveries have been followed since by a limited number of samples positive for any *Marburgvirus* that causes disease in humans. Despite decades of outbreaks in humans and extensive animal surveillance efforts, including unsuccessful attempts prior to the early 2000s, there is no approved vaccine or therapeutic that specifically targets MVD, and no MCM has progressed past Phase I trials (Figure 3C). Concerted pre-clinical and clinical-stage MCM development followed the 2004-2005 Angola outbreak, with a plurality of DNA and viral vector vaccine candidates designed specifically using viral isolates from infections during this outbreak (Supplementary Information). Research for vaccine and drug candidates has progressed during and after subsequent human outbreaks and without any distinctive relationship to viral discovery in animal hosts, even upon the discovery of a novel strain of Marburg virus through PREDICT.

EVD outbreaks, animal surveillance, and MCM development present similar patterns. MCM development primarily has followed the historic 2013-2016 West Africa epidemic, and animal surveillance has found limited success detecting any *Ebolavirus* in samples (Figure 4). The Zaire species has caused a plurality of EVD outbreaks since the first documented cases in 1976 (Figure 4A). Several investigations to discern outbreak origins have offered inconclusive evidence as to the source of disease or implicated flare-ups of undetected human-to-human transmission. In two recent notable instances of 2021 outbreaks in Guinea and the Democratic Republic of the Congo, epidemiological investigations traced the outbreaks to persistent viral infections and traced non-spillover-related transmission chains, calling into question the degree to which zoonotic spillover drives EVD outbreaks (Supplementary Information). Another study of the historic 2013-2016 epidemic raises questions about the hypothesis that the outbreak originated through a spillover event from a bat to a child near a hollow tree in the Guinean village of Meliandou, especially with no viral genomic material recovered near this site.^56^ Extended interviews with villagers have suggested instead that a persistently infected survivor from Sierra Leone in close contact with the child and his family may have first transmitted the virus.^57^

**Figure 4:**
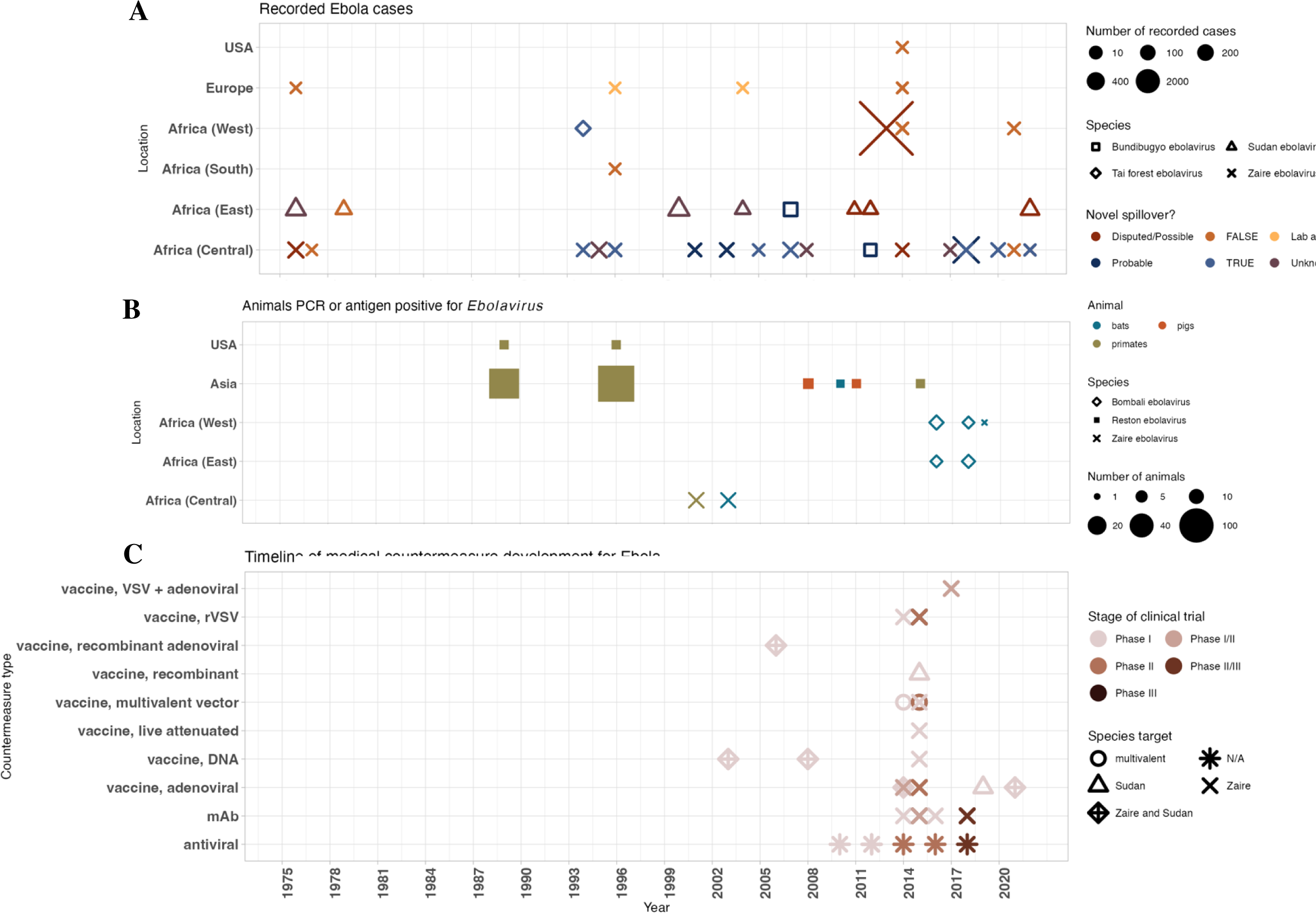
Ebola outbreaks, animal surveillance, and MCM development. A) Timeline of EVD outbreaks by species, assessment of novel spillover event as source of outbreak, and location. B) Timeline of when and where samples from animals were positive for an Ebolavirus by polymerase chain reaction or antigen test. C) Timeline of clinical trials by type of medical countermeasure specifically targeting an Ebolavirus.

Simultaneously, animal viral discovery has done little to prospectively identify novel sources of spillover for EVD. In 2016, PREDICT detected a novel species of *Ebolavirus*, which was named the Bombali virus and per one preliminary study could mediate entry into human cells (Figure 4B).^58^ However, more recent *in vivo* and *in vitro* studies suggest that *Bombali ebolavirus* has low pathogenic potential in humans, and it has not caused a documented outbreak in humans.^59, 60^ Reston ebolavirus is not known to cause disease in humans but has been detected more frequently than any other *Ebolavirus*, often in diseased non-human primates in laboratory settings. No viral prospecting efforts have successfully isolated live virus from any putative reservoir species. *Taï forest ebolavirus* was discovered when a researcher was infected during an investigation of disease in chimpanzees, likely while conducting a necropsy, but has not caused any documented outbreaks since (Supplementary Information). Furthermore, no Bombali-specific MCM development has progressed to a pre-clinical phase, and no MCM has specific regulatory approval for use against Sudan ebolavirus (Figure 4C). MCM development against *Ebolavirus* largely has followed the 2013-2016 epidemic and focused on *Zaire ebolavirus*, such that no approved MCMs were available to aid in responses for recent resurgences of *Sudan ebolavirus*.

## Discussion

This Analysis finds that viral prospecting and animal surveillance writ large have played a limited role in advancing preparedness and response for viral infectious diseases in humans as evaluated primarily through MCM development. These patterns suggest that viral prospecting does not address a rate-limiting step for preparedness efforts, especially when considered in conjunction with vast estimates of viral diversity and few parameters to guide high-yield searches for viruses of consequence for human disease. Our findings raise questions about whether viral prospecting is feasible, necessary, or sufficient as a means of predicting disease emergence and driving MCM development for emerging infectious diseases.

On feasibility, we note that searches for viruses in animals generally have been inadequate for pre-emptive and, to a lesser extent, post-hoc identification of viral threats to humans. Prospecting efforts in animals to date have had limited success in identifying novel viruses likely to pose substantial threats of outbreaks in humans. Efforts to definitively establish animal reservoirs for known human pathogens have had notable failures so far, most notably for ebolaviruses. Animal surveillance has achieved limited success in isolating from wildlife, especially bats, PCR or antigen-positive samples of viruses that cause disease in humans. It has only isolated one novel Ebola or Marburg species (Bombali ebolavirus), which seems as-yet unable to cause outbreaks in humans — as with other novel filoviruses discovered through viral prospecting^61, 62^ — and has received scant attention as a target for MCM development.^63, 64^

On sufficiency, we find for the 11 viruses to our knowledge discovered in a zoonotic host before causing documented clusters of cases in humans that there have been few differences in capacity for preparedness and response relative to other threats first discovered during a human outbreak, as well as no identifiable advances in preparedness between discovery in an animal and subsequent human outbreaks. Most of these viruses do not (yet) cause notable outbreaks in humans. Three viruses have caused several significant outbreaks, and MCM development against these three has not progressed differently from viral threats discovered in other ways. We note that while mosquito-based surveillance, especially for yellow fever, has identified several viruses with the ability to cause disease in humans and enabled various ecological and virological studies during and prior to outbreaks, systematic animal surveillance efforts in wildlife or domestic animals — the focus of most viral prospecting efforts today — have found comparatively limited success.

On necessity, there are still no vaccines approved for use in humans against any virus in 10 of 26 virus families with at least one virus known to infect humans, and the majority of disease outbreaks in humans involve known human pathogens or close relatives of known human pathogens. Even within the 16 of 26 virus families that contain at least one virus with an approved vaccine, several known threats (e.g.., Zika, West Nile, Nipah, Lassa) with demand for a vaccine do not have a countermeasure approved for widespread use. This undermines the hypothesis that a lack of candidate pathogens known prior to outbreaks is a limiting factor in our ability to develop MCMs and advance preparedness. On the contrary, most priority viruses for current MCM development efforts are pathogens that were discovered several decades ago but remain “high-value” targets with a lack of effective interventions, for reasons ranging from technical challenges to market failures.^65^ Larger regional outbreaks have instead been the catalyzing force for accelerated R&D, especially with vaccines. These data do not account for early-stage research for vaccine candidates often funded by various national funding agencies. However, they reflect the degree to which R&D efforts have not exhausted the work required to translate from known viral threats to approved vaccines against them.

Our analyses of filoviruses point at a larger body of research regarding viruses with known zoonotic hosts or assumed reservoirs that sometimes reemerge due to undetected or new modes of human-to-human transmission rather than new spillovers.^66, 67, 68, 69, 70, 71, 72, 73^ This raises questions about the degree to which outbreak frequency is increasing relative to public health capacity to detect outbreaks and coordination between local health efforts or institutions and international public health bodies. The case for being able to prospectively identify the source of a possible future outbreak from an animal reservoir to advance preparedness is relatedly tenuous, in light of notable difficulties associated with retrospectively discerning the origins of known outbreaks. Surveillance in animals with the goal of finding viruses with a high risk for spillover presumes that we have valid and reliable predictors of what viruses are likely to spill over and from where. The fact that we often cannot definitively isolate the putative virological source of an outbreak suggests that we do not have the data to generate such an account. Repeated spillovers of known viruses and flare-ups of human transmission that was previously undetected each contribute to the burden of disease emergence in a way that viral prospecting cannot significantly address. The filoviruses case study and our analyses more broadly instead suggest that existing disease in humans offers the best frameworks for preparedness and proxy for future disease widespread events in humans.

These analyses have some limitations, including a exclusive focus on viral pathogens (not bacterial or other zoonoses). First, filoviruses present one possible “best-case” scenario for MCM development, given extensive attention as a biosecurity concern. They do not pose the same challenges of respiratory transmission and high mutation rates that might more strongly motivate discovery efforts for coronaviruses or orthomyxoviruses (influenza). Second, our analysis mainly addresses a very specific endpoint of regulatory approval or lack thereof for the use of a vaccine in human populations, but it neither evaluates general capacity for scaled-up, widespread MCM manufacturing and equitable access nor accounts for veterinary vaccine development. Third, we do not make any explicit cost-benefit or risk-reward assessments. Finally, this analysis considers neither capacity-building and behavioral interventions nor research characterizing mechanisms such as viral entry or replication as a result of viral prospecting. We also note that the characterization of viral diversity and detection of viral threats are two distinct goals implicated in virus prospecting projects, and our study focuses on the latter. It relatedly does not account for less tangible ways in which prior knowledge about viruses, general healthcare system strengthening, or non-pharmaceutical interventions have factored into outbreak responses but does not equate the lack of direct MCM development with a lack of value for virus prospecting.

However, one possible implication of our analysis is that capacity-building through other activities such as (i) the surveillance of endemic viruses or disease in humans, rather than animals, and (ii) the prevention of ongoing spillover of known viruses, such as Nipah, by measures to reduce human contact with reservoirs and materials they contaminate,^74^ might still advance similar characterization-style goals of basic research. Such an approach may also contribute more concretely and reliably to preparedness efforts against future known and unknown threats, whether through vaccine development projects or other such initiatives that require dedicated technical, political, and financial resources.

We additionally evaluate two major plausible responses to our data and conclusions. The first is that these results only underscore the need for more extensive viral prospecting and MCM development efforts by highlighting the shortcomings of limited resources and efforts thus far. We note, however, the demonstrated difficulty of robustly assessing the pathogenicity of all viruses discovered through such initiatives and the various bottlenecks associated with translating knowledge of a virus to a usable vaccine or therapeutic. Scaling viral prospecting would not address these translational gaps, which themselves suggest that technical deficits in preparedness lie more in R&D or health-related infrastructure rather than predictions about viruses to target. A second response is that increased animal surveillance would allow more work to manage and curb human-wildlife interactions to stop possible spillover events before they even occur, especially in developing countries and through practices such as bushmeat hunting.^75^ However, the realities of an unequal and globalized world complicate this focus as a simple or all-encompassing policy intervention. For example, bats are a source of subsistence food and income in some rural communities in Africa,^76^ while several internationally driven development projects have disrupted ecosystem dynamics in ways that have brought humans and animals closer together and exacerbated disease risk.^77^ On the other hand, intensive agricultural practices^10^ or various factory farming-related risks of spillover from animal hosts for countries like the United States have gone somewhat unattended, such that responses to recent zoonotic outbreaks such as H5N1 from cattle have suffered from a lack of coordinated infrastructure. To address risks of spillover, then, would involve addressing social and economic dimensions of the contexts in which disease emerges. On their own, efforts to discover novel viruses pre-emptively offer little by way of predicting the ways in which people will respond to disease threats or deem preparedness measures adequate, suggesting that prediction as a paradigm itself requires refinement to offer legitimate predictive “power” for future problems.^78^

Further analyses of animal surveillance and viral prospecting might develop case studies across different virus families to specify the conditions under which such efforts would be most beneficial or track publications over time directly linked to linked to discoveries from viral prospecting. Additional work might chart MCM development timelines across clinical trials and MCM type across and within each virus family or characterize sampling efforts by assessing genomic diversity of sequences from animal surveillance relative to known human infections.

In terms of interventions for preparedness and response such as surveillance and R&D, history suggests that prospecting for novel viruses is unlikely to find the next Disease X before it “finds us.” We suggest that public health and policy decision-makers consider what specific forms of information and coordination allow for more targeted efforts that matter to people’s lives in the face of infectious disease threats instead of merely populating a universalizable database of viral zoonoses. As others have proposed, more focused serological and viral surveillance for disease in humans who are in contact with wildlife or livestock might provide more effective proxies for emerging disease risks and a clearer picture of disease burden while furthering knowledge of viral ecology.^29, 79^ Organizations like CEPI were created to address gaps in R&D efforts, especially in relation to market failures. Further efforts to advance clinical trials during outbreaks, including through innovative trial design and use of correlates of protections, and to develop MCMs that can be stockpiled prior to an outbreak would work to close gaps for known threats in ways that would build a stronger foundation for responses to yet-unknown viruses.

The lack of promised benefits from animal surveillance, alongside the extensive costs and possible risks referenced in the Introduction, point in favor of an argument for focusing resources elsewhere. These alternative directions offer other ways to expand capacity and further basic science — whether by improving MCM development efforts for known but under-characterized endemic threats of increasing epidemic concern, or strengthening local healthcare infrastructure to better ascertain and address the burden of diseases like Ebola and Marburg. Concerted attention to repeated and ongoing outbreaks in humans, especially in low and middle-income countries, would serve human health writ large and bolster preparedness for future global outbreaks in the event of increased spread. Mpox, the disease associated with monkeypox virus, offers an example of why such an approach merits further consideration. First, decades of increased disease incidence in parts of Africa went ignored internationally until a widespread global epidemic and PHEIC in 2022.^80^ Second, one recent study suggests that what was once thought to be spillover-induced spread of disease was in fact undetected human-to-human transmission.^81^ Third, in keeping with considerations around equity, as of April 2024 populations affected in Africa are yet to receive a dose of the smallpox-monkeypox vaccine.^82^ The fact that scientists discovered monkeypox in an animal prior to the first documented outbreak of the disease in humans has had little bearing on these challenges today.

In summary, viral prospecting in nonhuman animals does little to detect disease threats of consequence for human populations and has little to show in terms of advancing translational research for MCMs. Notwithstanding the value of acquiring further knowledge about viral diversity and expanding scientific capacity, narrow demonstrated benefits raise questions about whether other modes of preparedness might offer more suitable ways of achieving similar and further ends, without additional and inherent tradeoffs in cost, safety, or other domains. Closer attention to viral diseases in humans, whether emergent or known and often under-addressed, might instead lend itself to addressing inequalities and baseline capacity related to routine health needs across the world in ways that simultaneously strengthen preparedness and response against future emerging outbreaks.

## Methods

To broadly assess relationships between discovery in animals, disease outbreaks in humans, and preparedness through medical countermeasures across viral taxa, we began by referencing the ViralZone project^83^ (managed by the virus program of the Swiss-Prot group of the SIB Swiss Institute of Bioinformatics), International Committee on Taxonomy of Viruses (ICTV) lists from 2022,^84^ and a 2018 paper on Classification of Human Viruses.^53^ We first assessed the ViralZone list for viruses that were isolated in animals prior to any subsequent clusters of cases in humans, excluding viruses for which only singleton human cases have been reported to focus on pathogens of public health concern for potential outbreaks. We also used these lists to determine the first recorded outbreak of any virus from each family with at least one virus known to infect humans, either by conducting that search or systematically evaluating each virus known to infect humans in each family.

We approached the question of viral discovery once more by analyzing documented outbreaks in humans. We subset Carlson et al.’s Disease Outbreak News database^47^ to only include reports of viral diseases and then analyzed the geographic and virological distribution of emerging disease threats. We report frequency of DONs, noting that extended outbreaks are represented with more frequent and recurring weekly reports. As a result, our visualization of DONs offers a proxy for both the occurrence of a disease threat and the magnitude of it, which we chose over number of cases or disability-adjusted life years to reflect national and international public health institutions’ concerns regarding disease preparedness and response rather than epidemiological or economic statistics alone. We then referenced a WHO list of Public Health Emergency of International Concern (PHEIC) declarations and performed a similar analysis of disease origins.

Our analysis of priority pathogens drew from the 4 lists created by international and national bodies following the 2013-2016 Ebola epidemic, namely: WHO, NIAID (Category A), UKVN, and CEPI. We compiled priority pathogens across these lists to focus on viral threats and then conducted similar reviews to ascertain the circumstances in which each virus was first isolated. The figure excludes smallpox, which only the NIAID lists prioritizes and on biosecurity grounds, because the WHO declared the virus eradicated in 1980 and the first of several vaccines against the virus was developed in 1796.

We then used the virus families framework as described by Graham and Sullivan,^85^ among others, to evaluate the vaccine development landscape across virus families. We conducted a literature review to determine the year in which a vaccine was first approved for use in humans, primarily by a regulatory body — with the exception of rabies and smallpox, for which widespread vaccination preceded any regulatory approval. We coupled our evaluation of vaccine development with our analyses of when and how the first virus associated with each family was isolated, irrespective of the specific virus’ ability to infect humans. We focused on vaccines as opposed to other medical countermeasures because of our focus on public health approaches to preparedness over treatment in clinical settings, although we acknowledge the importance of therapeutic and diagnostic development as other relevant aspects for future consideration.

For our filoviruses case study, we based our literature search on information from the United States Centers for Disease Control and Prevention’s (CDC) timeline of EVD and MVD outbreaks over time. We used this timeline to then expand data collection to World Health Organizational and sub-regional organizations’ outbreak disease outbreak reports for additional epidemiological information. These sources were used to find key publications through PubMed and article citations to answer specific questions surrounding evidence for novel spillover and general outbreak origins, the context within which an outbreak began, and the demographics within which the virus spread. Case counts for each outbreak are based on laboratory-confirmed and suspected cases, where possible. In ascertaining whether an outbreak was caused by a novel spillover event, we used the following schematic: 1) *True* if both genomic and epidemiological evidence supported a novel spillover event, given availability of genomic data; 2) *Probable* if relatively conclusive genomic or epidemiological evidence but lacking evidence of the other form (provided both methods were available) or some other acknowledgement of uncertainty; 3) *Disputed/Possible* with competing explanations and unresolved debates within the literature; 4) *Unknown* if there was no evidence to whether a novel spillover event or existing circulating infection could be identified as the source of the outbreak or if literature generally states that the origins of a particular outbreak are unknown; 5) *False* if both genomic and epidemiological evidence identified existing and circulating infections in humans as the outbreak source. We conducted literature review for studies that found samples positive for *Ebolavirus* or *Marburgvirus* by polymerase chain reaction or antigen test to characterize animal host discovery over time, corroborated by GenBank searches for sequencing data. We did not include studies that reported seropositivity for either filovirus because this analysis focuses on surveillance for viruses to further understandings of potential pathogens rather than presence or absence of known viruses that elucidate, for instance, traces of infection in animal hosts. These criteria exclude several studies in which animal hosts are seropositive for a filovirus but none are positive by a molecular test for the presence of virus. For similar reasons, we did not include the many studies that describe efforts that failed to detect any sample that was positive for an *Ebolavirus* or *Marburgvirus*. Finally, we searched PubMed for reviews of EVD and MVD countermeasure development using the search terms “ebola,” “marburg,” and “filovirus” with “countermeasures,” “vaccine,” “antibody,” and “antiviral.” These reviews guided the construction of a set of relevant MCMs of interest, which then informed a further literature search to ascertain the timeline of clinical development (e.g., not basic research) for these countermeasures. The lack of any clinical trial beyond Phase I for MVD motivated further characterization of preclinical research against marburgviruses. In parallel, the search terms “ebola,” “marburg,” and “filovirus” were used on ClinicalTrials.gov to determine the progression of various countermeasures through clinical trials. Our dataset primarily includes the first instance of a clinical trial in a particular phase for a particular vaccine or drug candidate and is not comprehensive as to include all trials for comparing different dosage regimes or studying efficacy in different populations, for example.

## Data Availability

Data used for this analysis are available in Supplementary Information.

## Abbreviations

CEPI: Coalition for Epidemic Preparedness Innovations
DON: Disease Outbreak News
EVD: Ebola virus disease
MCM: medical countermeasure
MVD: Marburg virus disease
NIAID: National Institute of Allergy and Infectious Diseases
PHEIC: Public Health Emergency of International Concern
UKVN: UK Vaccines Research and Development Network
USAID: United States Agency for International Development
WHO: World Health Organization

## End Notes

Supplementary Information is available for this paper.

## Author Contributions

M.L. and A.V.A. conceived of the study. A.V.A. developed and carried out data collection and analyses. All authors contributed to writing the manuscript.

The authors thank Erin Mordecai for her comments on a later-stage version of this manuscript and multiple participants at the Ecology and Evolution of Infectious Diseases Conference (Stanford University, 2024) for comments on an earlier version of this work.

