## Supplementary material for "What is the relationship between viral prospecting in animals and medical countermeasure development?": 0 Aatresh and Lipsitch Viral Prospecting Supplement.docx

Table of Contents

Table 1 supplementary references……………………………………………………………...…2

Figure 1 supplementary table 1 and references…………………………………………………...4

Table 2 supplementary references………………………………………………………………...6

Figure 2 supplementary table 2 and references…………………………………………………...7

Table 3 supplementary references……………………………………………………………….11

Figures 3 and 4 information about supporting data and references…………………………...... 17

***Table 1: Viruses first discovered in animals before causing an outbreak in humans***

| **Virus** | **References** |
| --- | --- |
| Barmah Forest | 1 |
| Bunyamwera | 2, 3 |
| Eastern equine encephalitis | 4 |
| Monkeypox | 5 |
| Ngari | 2, 6 |
| Puumala | 7 |
| Rift Valley Fever | 8 |
| Semliki Forest | 9, 10, 11 |
| Sindbis | 12, 13 |
| Venezuelan equine encephalitis | 14 |
| Zika | 15 |

***Figure 1: Geographic and viral distribution of WHO DON reports (1996-2019)***

Supplementary Table 1: Viral disease events in DON database

| **Disease** | **PathogenType** |
| --- | --- |
| Anthrax |  |
| Botulism |  |
| Buffalopox | Virus |
| Chikungunya | Virus |
| Cholera |  |
| Coccidioidomycosis | |
| Crimean-Congo haemorrhagic fever | Virus |
| Dengue fever | Virus |
| Diptheria |  |
| Dysentery |  |
| E. coli |  |
| Ebola virus | Virus |
| Elizabethkingia anophelis | |
| Enterovirus | Virus |
| Gonorrhea |  |
| Guillain-Barre syndrome | |
| Hand, foot, and mouth disease | Virus |
| Hantavirus | Virus |
| Hemolytic uremic syndrome | |
| Hepatitis A | Virus |
| Hepatitis E | Virus |
| Human coronavirus OC43 | Virus |
| Human immunodeficiency virus | Virus |
| Influenza A | Virus |
| Japanese encephalitis | Virus |
| Lassa fever | Virus |
| Legionellosis | |
| Leishmaniasis | |
| Leptospirosis | |
| Listeriosis |  |
| Lujo mammarenavirus | Virus |
| Malaria |  |
| Marburg fever | Virus |
| Measles | Virus |
| Meningococcal disease | |
| MERS-CoV | Virus |
| Monkeypox | Virus |
| Nipah virus | Virus |
| O'nyong-nyong fever | Virus |
| Oropouche fever | Virus |
| Pertussis |  |
| Plague |  |
| Polio | Virus |
| Pseudomonas aeruginosa | |
| Rabies | Virus |
| Rift Valley fever | Virus |
| Salmonella enterica | |
| SARS-CoV | Virus |
| Smallpox | Virus |
| St. Louis encephalitis | Virus |
| Staphylococcus | |
| Streptococcus suis | |
| Syndromic: cardiovascular | |
| Syndromic: diarrhoeal | |
| Syndromic: gastrointestinal | |
| Syndromic: haemorrhagic | |
| Syndromic: neurological | |
| Syndromic: respiratory | |
| Tick-borne relapsing fever | |
| Toxicity: bromide poisoning | |
| Toxicity: lead poisoning | |
| Toxicity: miscellaneous | |
| Transmissible spongiform encephalopathy | |
| Tuberculosis | |
| Tularemia |  |
| Typhoid |  |
| Typhus |  |
| Unspecified |  |
| Venezuelan equine encephalitis | Virus |
| West Nile virus | Virus |
| Yellow fever | Virus |
| Zika virus disease | Virus |

***Table 2: WHO PHEIC declarations by viral disease and discovery status***

| **Year** | **Viral Disease** | **Novel Pathogen?** | **Notes** | **References** |
| --- | --- | --- | --- | --- |
| 2009 | H1N1 | TRUE |  | 1, 2 |
| 2014 | Ebola |  |  | 1, 3 |
| 2014 | Polio |  | PHEIC is ongoing | 1, 4 |
| 2016 | Zika |  |  | 1, 5 |
| 2018 | Ebola |  |  | 1, 6 |
| 2020 | COVID-19 | TRUE |  | 1, 7 |
| 2022 | mpox |  |  | 8, 9 |

| **Viral Pathogen/**  **Family** | **List** | **Year Isolated** | **How Discovered?** | **Zoonotic Transmission** | **Detection Detail** | **Notes** | **References** |
| --- | --- | --- | --- | --- | --- | --- | --- |
| MERS | CEPI, WHO, UK | 2012 | patient | TRUE | first human outbreak |  | 1, 2 |
| Lassa Fever | CEPI, UK, NIAID | 1972 | animal | TRUE | during human outbreak |  | 3 |
| Nipah | CEPI, UK | 1999 | patient | TRUE | first human outbreak |  | 4 |
| Rift Valley Fever | CEPI, WHO, UK | 1931 | animal | TRUE | pre human outbreak | Until 1975, RVF was regarded as an African, animal disease. Human cases were rare and with mild clinical manifestations. | 5 |
| Chikungunya | CEPI, UK | 1953 | patient | *vector-borne | during human outbreak |  | 6 |
| Ebola | CEPI, WHO, UK, NIAID | 1976 | patient | TRUE | first human outbreak |  | 7 |
| Marburg | WHO, UK, NIAID | 1967 | patient | TRUE | first human outbreak | in Europe, non endemic | 8 |
| COVID-19 | WHO | 2020 | patient | TRUE | first human outbreak |  | 9 |
| CCHF | WHO, UK | 1956 | patient | *vector-borne | during human outbreak | Disease Crimean Peninsula. Virus isolated Later Congo Basin. | 10, 11 |
| SARS | WHO | 2003 | patient | TRUE | first human outbreak |  | 12 |
| Henipaviruses | WHO | 1994 | animal + patient | TRUE | during human outbreak | Hendra virus | 13 |
| Zika | WHO, UK | 1947 | animal | *vector-borne | pre human outbreak | 1947 mosquitoes, 1952 humans. Found through yellow fever surveillance. First proof of human disease 1964. | 14 |
| Dengue | UK, NIAID | 1943 | patient | *vector-borne | during human outbreak | NIAID focuses on flaviviruses at the family level. | 15 |
| Hantaviruses | UK, NIAID | 1978 | animal | TRUE | post human outbreak | NIAID focuses on bunyaviruses at the family level, of which hantaviruses are one example. | 16 |
| Smallpox | NIAID | 1796 | patient | FALSE | during human outbreak | vaccine in 1796, Edward Jenner. Excluded from figure because i) existing vaccine with regulatory approval ii) disease eradicated iii) biosecurity concern specific to NIAID | 17 |

***Table 3: Vaccine development and viral discovery across virus families***

| **Family** | **References** |
| --- | --- |
| Adenoviridae | 1, 2, 3 |
| Anelloviridae | 4, 5 |
| Arenaviridae | 6, 7, 8, 9 |
| Astroviridae | 10, 11, 12 |
| Bornaviridae | 13, 14, 15, 16 |
| Bunyaviridae | 17, 18 |
| Caliciviridae | 19, 20, 21 |
| Coronaviridae | 22 |
| Filoviridae | 8, 23 |
| Flaviviridae | 24, 25, 26 |
| Hepadnaviridae | 27, 28 |
| Hepeviridae | 29, 30 |
| Herpesviridae | 31 |
| Orthomyxoviridae | 32 |
| Papillomaviridae | 33, 34, 35, 36, 37 |
| Paramyxoviridae | 38, 39, 40 |
| Parvoviridae | 41, 42, 43 |
| Picobirnaviridae | 44 |
| Picornaviridae | 45, 46, 47 |
| Pneumoviridae | 48, 49, 50 |
| Polyomaviridae | 51, 52, 53 |
| Poxviridae | 54, 55, 56 |
| Reoviridae | 57, 58, 59, 60 |
| Retroviridae | 61, 62 |
| Rhabdoviridae | 63, 64, 65 |
| Togaviridae | 66, 67, 68, 69, 70 |

1. Centers for Disease Control and Prevention. Adenovirus Vaccine Information Statement. *Vaccine Information Statements* https://www.cdc.gov/vaccines/hcp/vis/vis-statements/adenovirus.html (2020).
2. Soares, J. M. USAMMDA Seeks Refresh of Adenovirus Vaccine. *www.army.mil* https://www.army.mil/article/183287/usammda_seeks_refresh_of_adenovirus_vaccine (2017).
3. Robinson, C. M., Seto, D., Jones, M. S., Dyer, D. W. & Chodosh, J. Molecular evolution of human species D adenoviruses. *Infect Genet Evol* **11**, 1208–1217 (2011).
4. Harnessing the Untapped Potential of Anelloviruses. *Flagship Pioneering* https://www.flagshippioneering.com/stories/harnessing-the-untapped-potential-of-anelloviruses (2022).
5. Spezia, P. G. *et al.* TTV and other anelloviruses: The astonishingly wide spread of a viral infection. *Asp Mol Med* **1**, None (2023).
6. Carnec, X. *et al.* A Vaccine Platform against Arenaviruses Based on a Recombinant Hyperattenuated Mopeia Virus Expressing Heterologous Glycoproteins. *J Virol*  **92**, e02230-17 (2018).
7. Gowen, B. B. *et al.* Second-Generation Live-Attenuated Candid#1 Vaccine Virus Resists Reversion and Protects against Lethal Junín Virus Infection in Guinea Pigs. *J Virol* **95**, e0039721 (2021).
8. CDC. About Viral Hemorrhagic Fevers. *Viral Hemorrhagic Fevers (VHFs)* https://www.cdc.gov/viral-hemorrhagic-fevers/about/index.html (2024).
9. Johnson, D. M. *et al.* Bivalent Junin & Machupo experimental vaccine based on alphavirus RNA replicon vector. *Vaccine* **38**, 2949–2959 (2020).
10. Bosch, A., Pintó, R. M. & Guix, S. Human Astroviruses. *Clin Microbiol Rev* **27**, 1048–1074 (2014).
11. Human Astrovirus Vaccine. *Creative Biolabs* https://www.creative-biolabs.com/vaccine/human-astrovirus-vaccine.htm.
12. Bidokhti, M. R. M. *et al.* Immunogenicity and Efficacy Evaluation of Subunit Astrovirus Vaccines. *Vaccines (Basel)***7**, 79 (2019).
13. Dürrwald, R. *et al.* Vaccination against Borna Disease: Overview, Vaccine Virus Characterization and Investigation of Live and Inactivated Vaccines. *Viruses* **14**, 2706 (2022).
14. Honda, T. Relaunching human bornavirus research from encephalitis cases with unclear cause. *The Lancet Infectious Diseases* **20**, 389–391 (2020).
15. Bauswein, M. *et al.* Human Infections with Borna Disease Virus 1 (BoDV-1) Primarily Lead to Severe Encephalitis: Further Evidence from the Seroepidemiological BoSOT Study in an Endemic Region in Southern Germany. *Viruses* **15**, 188 (2023).
16. Tizard, I., Ball, J., Stoica, G. & Payne, S. The pathogenesis of bornaviral diseases in mammals. *Animal Health Research Reviews* **17**, 92–109 (2016).
17. Ikegami, T. & Makino, S. Rift Valley fever vaccines. *Vaccine* **27S4**, D69–D72 (2009).
18. Wichgers Schreur, P. J., Bird, B. H., Ikegami, T., Bermúdez-Méndez, E. & Kortekaas, J. Perspectives of Next-Generation Live-Attenuated Rift Valley Fever Vaccines for Animal and Human Use. *Vaccines (Basel)* **11**, 707 (2023).
19. Feline Calicivirus. *Cornell University College of Veterinary Medicine* https://www.vet.cornell.edu/departments-centers-and-institutes/baker-institute/research-baker-institute/feline-calicivirus.
20. Tan, M. Norovirus Vaccines: Current Clinical Development and Challenges. *Pathogens* **10**, 1641 (2021).
21. Caliciviridae. *Encyclopedia of Virology (Third Edition)* https://www.sciencedirect.com/topics/medicine-and-dentistry/caliciviridae (2008).
22. Krammer, F. The role of vaccines in the COVID-19 pandemic: what have we learned? *Semin Immunopathol* **45**, 451–468 (2024).
23. First FDA-approved vaccine for the prevention of Ebola virus disease, marking a critical milestone in public health preparedness and response. *U.S. Food & Drug Administration* https://www.fda.gov/news-events/press-announcements/first-fda-approved-vaccine-prevention-ebola-virus-disease-marking-critical-milestone-public-health (2019).
24. Tien, S.-M. *et al.* Therapeutic efficacy of humanized monoclonal antibodies targeting dengue virus nonstructural protein 1 in the mouse model. *PLOS Pathogens* **18**, e1010469 (2022).
25. Norrby, E. Yellow fever and Max Theiler: the only Nobel Prize for a virus vaccine. *J Exp Med* **204**, 2779–2784 (2007).
26. Gianchecchi, E., Cianchi, V., Torelli, A. & Montomoli, E. Yellow Fever: Origin, Epidemiology, Preventive Strategies and Future Prospects. *Vaccines (Basel)* **10**, 372 (2022).
27. Hepadnaviridae. *ScienceDirect | International Journal of Infectious Diseases* https://www.sciencedirect.com/topics/medicine-and-dentistry/hepadnaviridae (2008).
28. CDC. Chapter 10: Hepatitis B. *Epidemiology and Prevention of Vaccine-Preventable Diseases* https://www.cdc.gov/pinkbook/hcp/table-of-contents/chapter-10-hepatitis-b.html (2024).
29. Kelly, A. G., Netzler, N. E. & White, P. A. Ancient recombination events and the origins of hepatitis E virus. *BMC Evolutionary Biology* **16**, 210 (2016).
30. Sridhar, S. *et al.* A Systematic Approach to Novel Virus Discovery in Emerging Infectious Disease Outbreaks. *J Mol Diagn* **17**, 230–241 (2015).
31. World Health Organization. Herpes Simplex Virus. *Immunization, Vaccines and Biologicals* https://www.who.int/teams/immunization-vaccines-and-biologicals/diseases/herpes-simplex-virus (2022).
32. History of influenza vaccination. *World Health Organization* https://www.who.int/news-room/spotlight/history-of-vaccination/history-of-influenza-vaccination.
33. CDC. HPV Vaccination. *Human Papillomavirus (HPV)* https://www.cdc.gov/hpv/vaccines/index.html (2024).
34. KFF. The HPV Vaccine: Access and Use in the U.S. *Women’s Health Policy* https://www.kff.org/womens-health-policy/fact-sheet/the-hpv-vaccine-access-and-use-in-the-u-s/ (2021).
35. Dürst, M., Gissmann, L., Ikenberg, H. & zur Hausen, H. A papillomavirus DNA from a cervical carcinoma and its prevalence in cancer biopsy samples from different geographic regions. *Proc Natl Acad Sci U S A* **80**, 3812–3815 (1983).
36. Watts, G. Harald zur Hausen. *The Lancet* **402**, 20 (2023).
37. The Nobel Prize in Physiology or Medicine 2008. *The Nobel Prize* https://www.nobelprize.org/prizes/medicine/2008/advanced-information/.
38. Hendriks, J. & Blume, S. Measles Vaccination Before the Measles-Mumps-Rubella Vaccine. *Am J Public Health* **103**, 1393–1401 (2013).
39. Almansour, I. Mumps Vaccines: Current Challenges and Future Prospects. *Front Microbiol* **11**, 1999 (2020).
40. Johnson, C. D. & Goodpasture, E. W. AN INVESTIGATION OF THE ETIOLOGY OF MUMPS. *J Exp Med* **59**, 1–19 (1934).
41. Bernstein, D. I. *et al.* Safety and Immunogenicity of a Candidate Parvovirus B19 Vaccine. *Vaccine* **29**, 7357–7363 (2011).
42. Pénzes, J. J. *et al.* Reorganizing the family Parvoviridae: a revised taxonomy independent of the canonical approach based on host association. *Arch Virol* **165**, 2133–2146 (2020).
43. Union of International Associations. Parvoviruses. *THE ENCYCLOPEDIA OF WORLD PROBLEMS & HUMAN POTENTIAL* http://encyclopedia.uia.org/en/problem/parvoviruses.
44. Wang, D. The enigma of picobirnaviruses: viruses of animals, fungi, or bacteria? *Curr Opin Virol* **54**, 101232 (2022).
45. Polio Vaccination. *CDC* https://www.cdc.gov/vaccines/vpd/polio/index.html (2022).
46. History of polio vaccination. *World Health Organization* https://www.who.int/news-room/spotlight/history-of-vaccination/history-of-polio-vaccination.
47. Berhanu, A. PICORNAVIRIDAE. https://web.stanford.edu/group/virus/picorna/2005/PICORNAVIRIDAE.htm.
48. CDC. RSV Vaccine Information Statement. https://www.cdc.gov/vaccines/hcp/vis/vis-statements/rsv.html (2023).
49. FDA Approves First Respiratory Syncytial Virus (RSV) Vaccine. https://www.fda.gov/news-events/press-announcements/fda-approves-first-respiratory-syncytial-virus-rsv-vaccine (2023).
50. Collins, P. L. & Graham, B. S. Viral and Host Factors in Human Respiratory Syncytial Virus Pathogenesis. *J Virol***82**, 2040–2055 (2008).
51. Peretti, A. *et al.* A multivalent polyomavirus vaccine elicits durable neutralizing antibody responses in macaques. *Vaccine* **41**, 1735–1742 (2023).
52. Neu, U., Stehle, T. & Atwood, W. J. The Polyomaviridae: Contributions of virus structure to our understanding of virus receptors and infectious entry. *Virology* **384**, 389–399 (2009).
53. IARC Working Group on the Evaluation of Carcinogenic Risks to Humans. INTRODUCTION TO POLYOMAVIRUSES. in *Malaria and Some Polyomaviruses (SV40, BK, JC, and Merkel Cell Viruses)* (International Agency for Research on Cancer, 2013).
54. Sánchez-Sampedro, L. *et al.* The Evolution of Poxvirus Vaccines. *Viruses* **7**, 1726–1803 (2015).
55. History of Smallpox | Smallpox. *CDC* https://www.cdc.gov/smallpox/history/history.html (2024).
56. Thèves, C., Biagini, P. & Crubézy, E. The rediscovery of smallpox. *Clinical Microbiology and Infection* **20**, 210–218 (2014).
57. Rotavirus Vaccination. *CDC* https://www.cdc.gov/vaccines/vpd/rotavirus/index.html (2023).
58. CDC. Chapter 19: Rotavirus. *Epidemiology and Prevention of Vaccine-Preventable Diseases*https://www.cdc.gov/pinkbook/hcp/table-of-contents/chapter-19-rotavirus.html (2024).
59. Kim, M. Naturally occurring reoviruses for human cancer therapy. *BMB Rep* **48**, 454–460 (2015).
60. Sabin, A. B. Reoviruses: A new group of respiratory and enteric viruses formerly classified as ECHO type 10 is described. *Science* **130**, 1387–1389 (1959).
61. Coffin, J. M., Hughes, S. H. & Varmus, H. E. A Brief Chronicle of Retrovirology. in *Retroviruses* (Cold Spring Harbor Laboratory Press, 1997).
62. Vahlne, A. A historical reflection on the discovery of human retroviruses. *Retrovirology* **6**, 40 (2009).
63. Hicks, D. J., Fooks, A. R. & Johnson, N. Developments in rabies vaccines. *Clin Exp Immunol* **169**, 199–204 (2012).
64. Fooks, A. R., Banyard, A. C. & Ertl, H. C. J. New human rabies vaccines in the pipeline. *Vaccine* **37**, A140–A145 (2019).
65. Rabies Vaccine Information Statement. *CDC* https://www.cdc.gov/vaccines/hcp/vis/vis-statements/rabies.html (2023).
66. About Rubella (German Measles) Vaccination. *CDC* https://www.cdc.gov/vaccines/vpd/rubella/index.html (2022).
67. Western Equine Encephalitis. *ScienceDirect* https://www.sciencedirect.com/topics/medicine-and-dentistry/western-equine-encephalitis.
68. CDC. Chapter 20: Rubella. *Epidemiology and Prevention of Vaccine-Preventable Diseases*https://www.cdc.gov/pinkbook/hcp/table-of-contents/chapter-20-rubella.html (2024).
69. History and Timeline. https://web.stanford.edu/group/virus/1999/jchow/histtime.html (1999).
70. Brinton, M. A. Replication of Flaviviruses. in *The Togaviridae and Flaviviridae* (eds. Schlesinger, S. & Schlesinger, M. J.) 327–374 (Springer New York, Boston, MA, 1986). doi:10.1007/978-1-4757-0785-4_11.

.xlsx files indicate sources for data in the following figures:

***Figure 3: Marburg outbreaks, animal surveillance, and MCM development***

***Figure 4: Ebola outbreaks, animal surveillance, and MCM development***
